# Carbimazole, methimazole and propylthiouracil: Use in women of childbearing age and exposed pregnancies in Germany

**DOI:** 10.1101/2025.02.26.25322848

**Authors:** Tania Schink, Maxim Frizler, Bianca Kollhorst, Ulrike Haug

**Affiliations:** Department of Clinical Epidemiology, Leibniz Institute for Prevention Research and Epidemiology – BIPS, Bremen, Germany; Federal Institute for Drugs and Medical Devices (BfArM); Department of Biometry and Data Management, Leibniz Institute for Prevention Research and Epidemiology – BIPS, Bremen, Germany; Faculty of Human and Health Sciences, University of Bremen, Bremen, Germany

**Keywords:** Antithyroid drugs, carbimazole, methimazole, propylthiouracil, pregnancy

## Abstract

**Background:** Hyperthyroidism during pregnancy is associated with maternal, obstetrical and fetal complications. Antithyroid drugs (ATD) including carbimazole (CMZ), methimazole (MMI) and propylthiouracil (PTU) are the main pharmacotherapy for hyperthyroidism. Exposure to CMZ and MMI during the first trimester was associated with birth defects, while PTU is assumed to be the safer alternative.

**Objective:** To calculate the prescription prevalences of ATD in women of childbearing age over time and to describe pregnancies occurring after or during ATD use.

**Methods:** Using the GePaRD database (claims data; 20% of the German population), we conducted year-wise cross-sectional studies for the period 2004-2020 to calculate prescription prevalences of CMZ, MMI and PTU in females aged 13-49 years. In longitudinal analyses, we included all women with any ATD dispensing between 2005 and 2020 aged 13- 49 years at the first dispensing. We identified pregnancies occurring in this cohort and described ATD use before and during pregnancy.

**Results:** The age-standardized prescription prevalence of ATDs decreased by 32.1% between 2004 (2.71 per 1,000) and 2020 (1.84 per 1,000). This decrease was attributable to CMZ (2004: 1.40 per 1,000; 2020: 0.76 per 1,000; relative decrease: 45.7%) and MMI (2004: 1.25 per 1,000; 2020: 0.99 per 1,000; relative decrease: 20.8%). In the cohort including 9,723 women, 13,586 pregnancies were observed, of which 67% (n=9,140) occurred after ATD use. In 16.2% of the pregnancies (n=2,203), ATD use overlapped with pregnancy onset. The proportion exposed to CMZ/MMI at pregnancy onset decreased from 30.7% in 2005 to 10.9% in 2020. In 16.5% of pregnancies (n=2,243), ATD use was started during pregnancy.

**Conclusion:** The prescription prevalence of ATD overall and specifically of CMZ/MMI in girls and women of childbearing age decreased between 2005 and 2020 in Germany. The decrease in exposure to CMZ/MMI at pregnancy onset indicate that physicians became increasingly aware of the potential risks of CMZ/MMI to the unborn child.

## Introduction

Hyperthyroidism during pregnancy, which is mainly caused by Graves’ disease, is associated with maternal, obstetrical and fetal complications. Therefore, treatment of hyperthyroidism should ideally be completed before pregnancy onset.(1) Antithyroid drugs (ATDs), i.e., methimazole (MMI) and its prodrug carbimazole (CMZ) as well as propylthiouracil (PTU), play an important role in the treatment of hyperthyroidism. Results of observational studies, however, suggested an increased risk of birth defects for CMZ/MMI, while PTU is assumed to be the safer alternative during pregnancy, even though it can induce liver injury in the mother.(2, 3)

In November 2018, the Pharmacovigilance Risk Assessment Committee (PRAC) of the European Medicines Agency (EMA) recommended that women of childbearing potential should use effective contraceptive measures during treatment with CMZ/MMI. The PRAC also stated that hyperthyroidism in pregnant women should be adequately treated to prevent serious maternal and fetal complications and that the use of CMZ/MMI in pregnant women should be based on an individual benefit/risk assessment.(4) In addition, the guideline of the European Thyroid Association published in 2018 recommended that women treated with CMZ/MMI should be switched to PTU when planning a pregnancy and/or during the 1^st^ trimester of pregnancy. After gestational week 16, a switch from PTU to CMZ/MMI should be considered.(5)

To the best of our knowledge, there is no study investigating prescribing of ATDs in women of childbearing age and during pregnancy in Germany. Against this background, we aimed to determine the prescription prevalence of ATDs in women of childbearing age over time, to describe pregnancies occurring under ATDs and to examine switching patterns between CMZ/MMI and PTU before and during pregnancy in Germany.

## Methods

### Data Source

We used the German Pharmacoepidemiological Research Database (GePaRD) for this study, which is based on claims data from four statutory health insurance providers in Germany and currently includes information on approximately 25 million persons who have been insured with one of the participating providers since 2004 or later. In addition to demographic data, GePaRD contains information on drug dispensings as well as outpatient (i.e., from general practitioners and specialists) and inpatient services and diagnoses. Per data year, there is information on approximately 20% of the general population and all geographical regions of Germany are represented.(6)

### Prescription prevalence

We conducted year-wise cross-sectional analyses for the years 2004-2020 to assess the prescription prevalence of CMZ, MMI, and PTU as well as changes over time. For each calendar year, we included all girls and women in the numerator who had at least one dispensing of the respective medication, were aged between 13 and 49 years in the respective year, were insured on June 30 of that year and were residents in Germany. In the denominator, we included all girls and women aged between 13 and 49 years in the respective year, insured on June 30 of that year and with residence in Germany.

For each year, we calculated age-specific and age-standardized prescription prevalences. For age-standardization, we used the age distribution of the German female population on December 31, 2019.

### Pregnancies among women using ATDs

The study period for the inclusion of pregnancies started on January 1, 2005 and ended on December 31, 2020. First, we built a cohort of all women residing in Germany with at least one dispensing of an ATD between 13 and 49 years of age during the study period and at least one year of continuous insurance before the ATD dispensing leading to cohort entry. Cohort entry was defined as the date of the first ATD dispensing at which the above criteria were fulfilled. Cohort exit was defined as the first of the following dates: (i) end of insurance (i.e., death, end or interruption of insurance for more than 30 days), (ii) end of the year in which the woman turned 50, and (iii) end of the study period, i.e., December 31, 2020.

In this cohort, we identified all pregnancies overlapping with the time in the cohort, yielding the pregnancy cohort that was further analyzed. Pregnancies were identified using a validated outcome algorithm, which also provides information on the end date of the pregnancy.(7) Pregnancy onset could mostly be estimated using the expected delivery date, which is determined based on the last menstrual period (LMP) or early ultrasound.(8) In addition to the outcome algorithm, we also searched for pregnancies with no outcome recorded in claims data (e.g., miscarriages not requiring medical treatment, induced abortions not reimbursed by the statutory health insurance because there was no medical indication). To qualify for this category, there had to be at least one code indicating the expected delivery date and another indicator of a pregnancy (e.g., a pregnancy-related examination) within a plausible time interval after the estimated beginning of pregnancy. The date of the last pregnancy-related examination recorded in the data was assigned as the end of pregnancy in this category.

We defined ATD treatment episodes starting on the dispensing date. Each treatment episode started with the date of the respective dispensing and continued as long as there were further dispensings within the time covered by two times the number of DDDs. This approach takes into account potential lower doses of ATDs, i.e. it avoids underestimating the duration of the treatment episodes. Exposure during pregnancy was defined as an ATD treatment episode overlapping pregnancy onset or an ATD dispensing during pregnancy.

For the assessment of treatment patterns, we calculated the proportion of pregnancies exposed to ATDs (overall and on a substance level), the proportion of pregnancies with additional CMZ/MMI or PTU dispensing during pregnancy, the proportion with a switch from CMZ/MMI to PTU before or during pregnancy and from PTU to CMZ/MMI during pregnancy and assessed the timepoint of the switches relative to pregnancy onset.

### Ethics statement

In Germany, the utilization of health insurance data for scientific research is regulated by the Code of Social Law. All involved health insurance providers as well as the German Federal Office for Social Security and the Senator for Health, Women and Consumer Protection in Bremen as their responsible authorities approved the use of GePaRD data for this study. Informed consent for studies based on claims data is required by law unless obtaining consent appears unacceptable and would bias results, which was the case in this study. According to the Ethics Committee of the University of Bremen studies based on GePaRD are exempt from institutional review board review.

## Results

### Prescription prevalence

The age-standardized prescription prevalence of ATDs in girls and women of childbearing age decreased by about one third between 2004 and 2020 (2004: 2.71 per 1,000; 2020: 1.84 per 1,000). This overall decrease was driven by CMZ, for which the age-standardized prescription prevalence decreased by 45.7% (2004: 1.40 per 1,000; 2020: 0.76 per 1,000) as well as by MMI, for which the age-standardized prescription prevalence decreased by 20.8% (2004: 1.25 per 1,000; 2020: 0.99 per 1,000). During the same period, the age-standardized prescription prevalence of PTU was stable (Fig 1).

**Fig 1.**
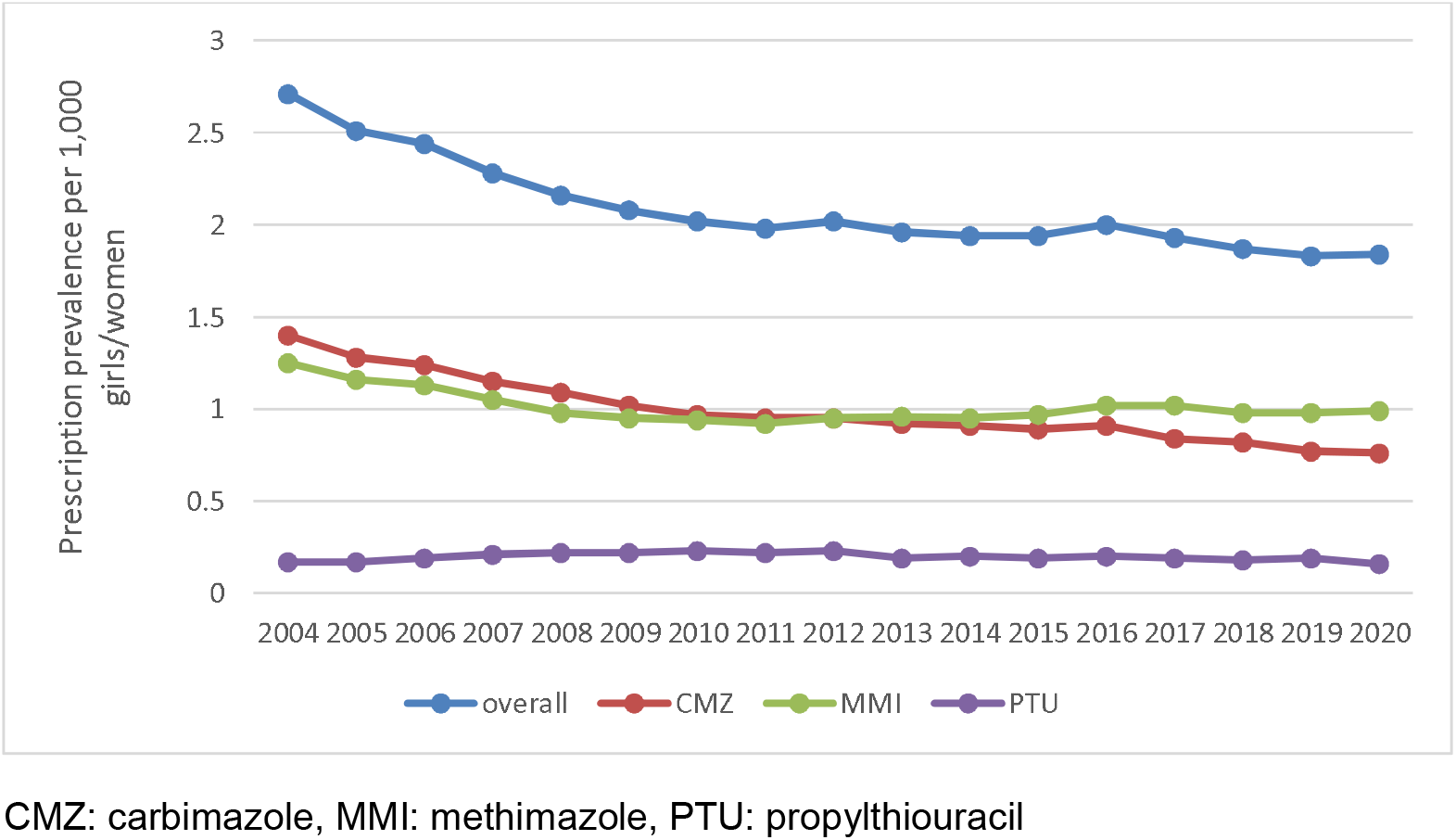
Age-standardized prescription prevalence of ATDs (overall and by substance) in girls and women of childbearing between 2004 and 2020.

The prescription prevalence of CMZ and MMI decreased from 2004 to 2020 in women older than 26 years, but remained stable in girls and women younger than 26 years. Regarding PTU, no trends over time were observed (Fig 2).

**Fig 2.**
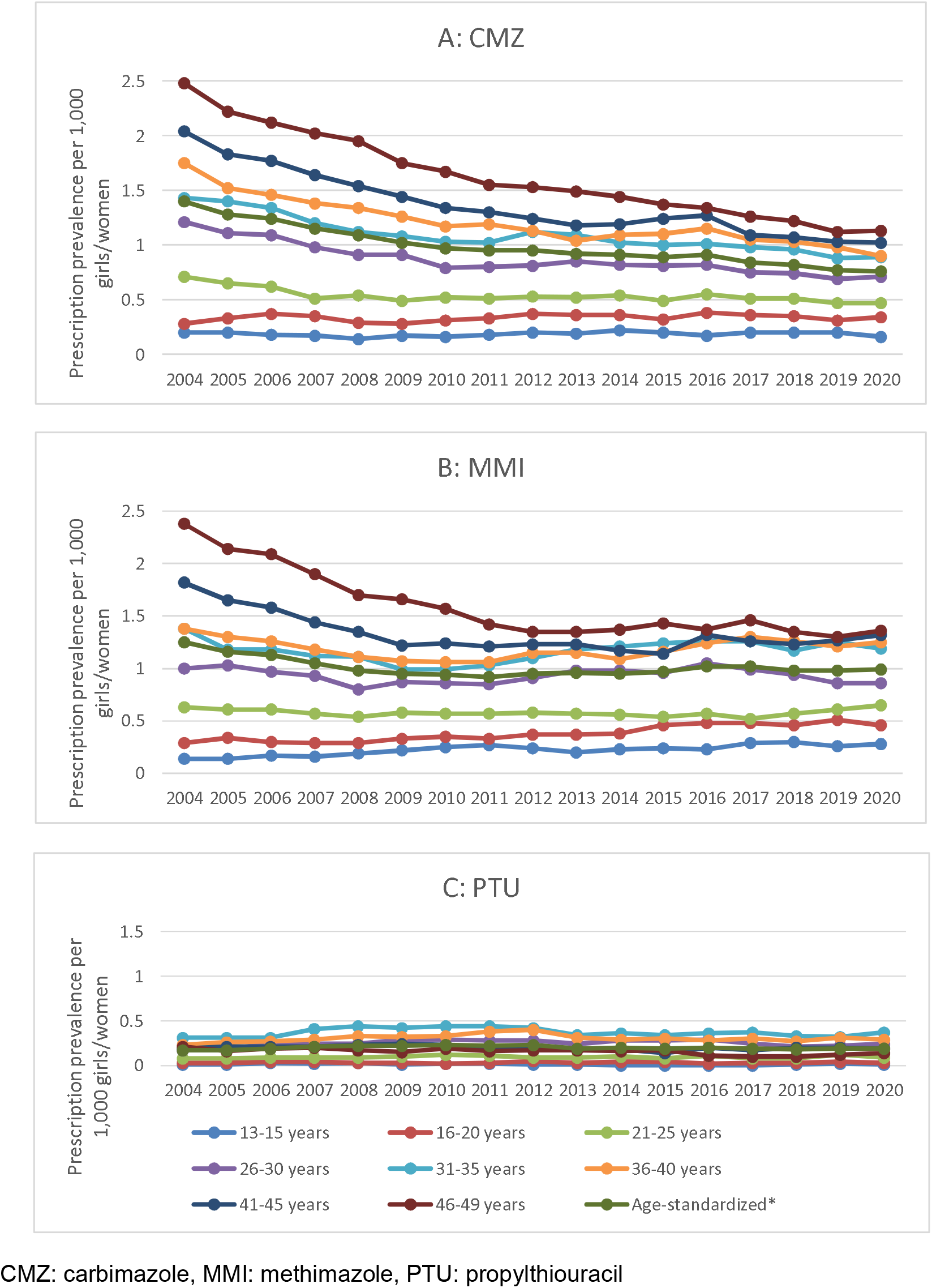
Age-specific prescription prevalence of ATDs (by substance) in girls and women of childbearing age between 2004 and 2020 (A: CMZ, B: MMI, C: PTU)

The prescription prevalence of CMZ and MMI increased with the age of the girls and women. For PTU, there was a different age pattern: the prescription prevalence was low in the youngest age groups, higher in the oldest age groups, and the highest prescription prevalence was observed in the age group 31-35 years (Fig 2).

### Pregnancies

In the cohort of 57,516 women with an ATD dispensing, 13,586 pregnancies were observed in 9,723 women (16.9%). The median age of women at pregnancy onset was 33 years (25^th^- 75^th^ percentile: 30-36 years).

In 67.3% of pregnancies (n=9,140), no ATDs were used, i.e., women were treated with ATDs before pregnancy, but there was neither an overlap between ATD treatment episodes and pregnancy onset nor an ATD dispensing during pregnancy. In 16.2% of pregnancies (n=2,203), the woman was on ATD medication at pregnancy onset. This proportion decreased from 33.6% in 2005 to 12.8% in 2020. This was mainly driven by the decrease in the proportion exposed to CMZ/MMI (30.7% in 2005; 10.9% in 2020). (Fig 3) In 16.5% of pregnancies (n=2,243), ATD medication was started during pregnancy.

**Fig 3.**
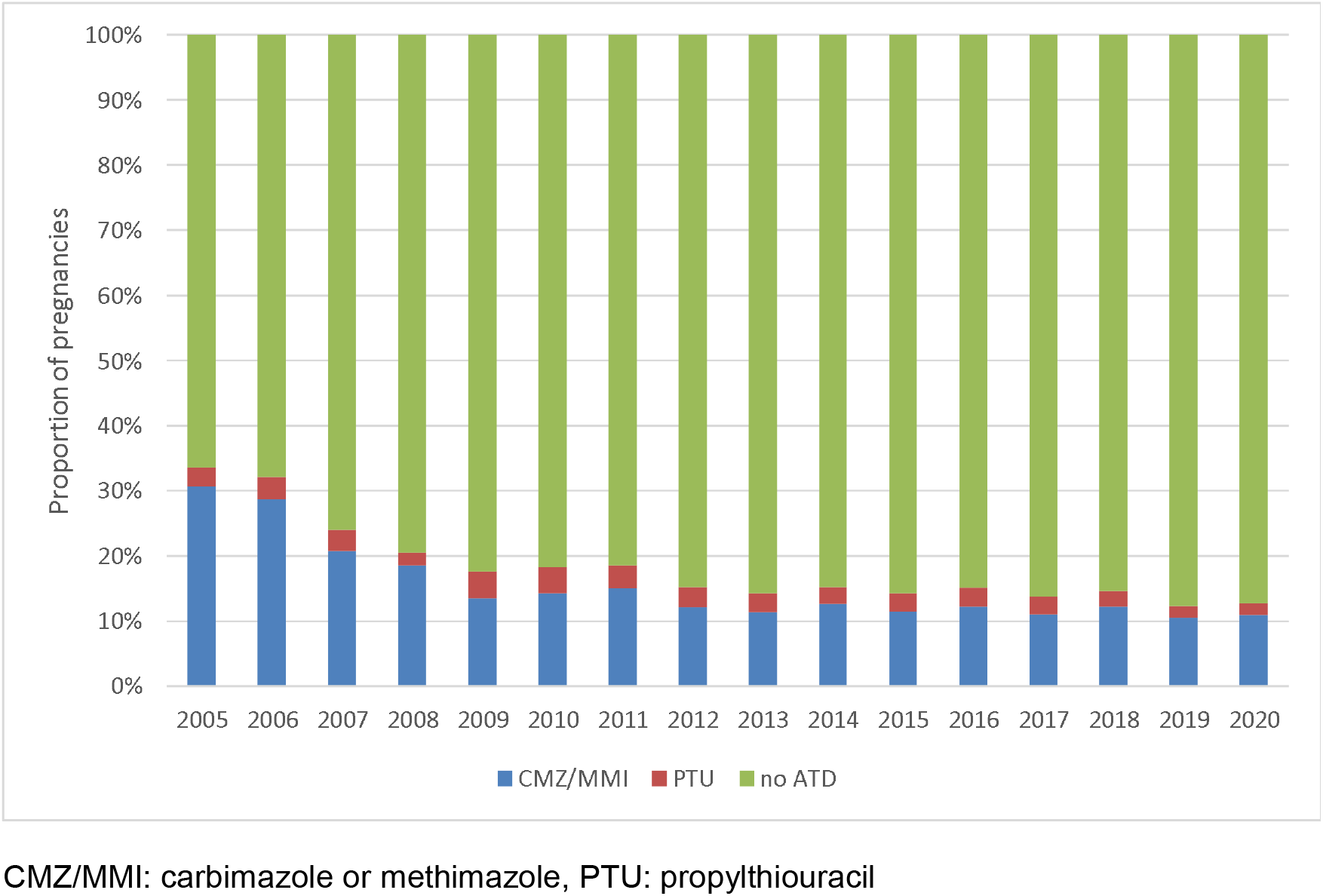
Pregnancies occurring in the cohort of ATD users: Distribution regarding ADT exposure at pregnancy onset.

Among the 2,203 pregnancies exposed to ATDs at pregnancy onset, 82.8% (n=1,842) were exposed to CMZ/MMI. This proportion was 91.4% in 2005 and 85.2% in 2020. The remaining pregnancies (n=379) were exposed to PTU.

#### CMZ/MMI at pregnancy onset

Among the 1,824 pregnancies exposed to CMZ/MMI at pregnancy onset, 36.2% (n=660) had at least one additional CMZ/MMI dispensing during pregnancy: in 83.3% of these pregnancies, there was a further dispensing during the first trimester, in 35.2% during the second trimester, and in 21.8% during the third trimester. The proportion of pregnancies in which CMZ/MMI was dispensed during the first trimester decreased between 2005 (41.9%) and 2020 (23.9%) (Fig 4).

**Fig 4.**
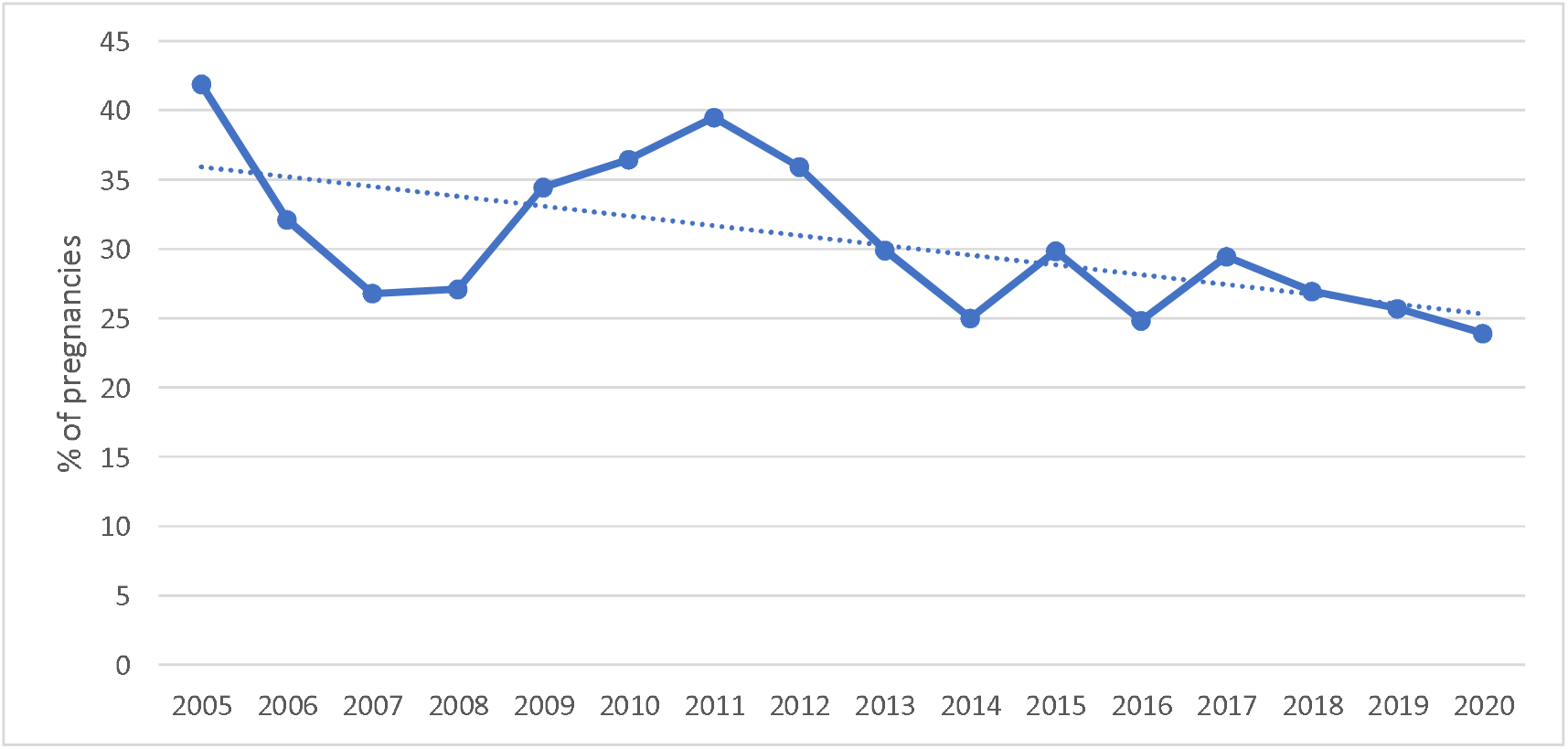
Exposure group “CMZ/MMI at pregnancy onset”: Proportion of pregnancies with additional CMZ/MMI dispensing during the first trimester.

A switch from CMZ/MMI to PTU during pregnancy was observed in 16.2% of the 1,824 pregnancies (n=296). In 65.2% of the switches, there was no additional CMZ/MMI dispensing before the switch. The switch from CMZ/MMI to PTU occurred in median 8 weeks after pregnancy onset (25^th^-75^th^ percentile: 6-10 weeks). The proportion of pregnancies with a switch from CMZ/MMI to PTU increased between 2005 (6.8%) and 2020 (23.9%) (Fig 5). About a quarter (23.3%) of switchers re-switched to CMZ/MMI during pregnancy. The re-switch occurred in median 17 (14-23) weeks after pregnancy onset and 10 (7-16) weeks after the switch from CMZ/MMI to PTU.

**Fig 5.**
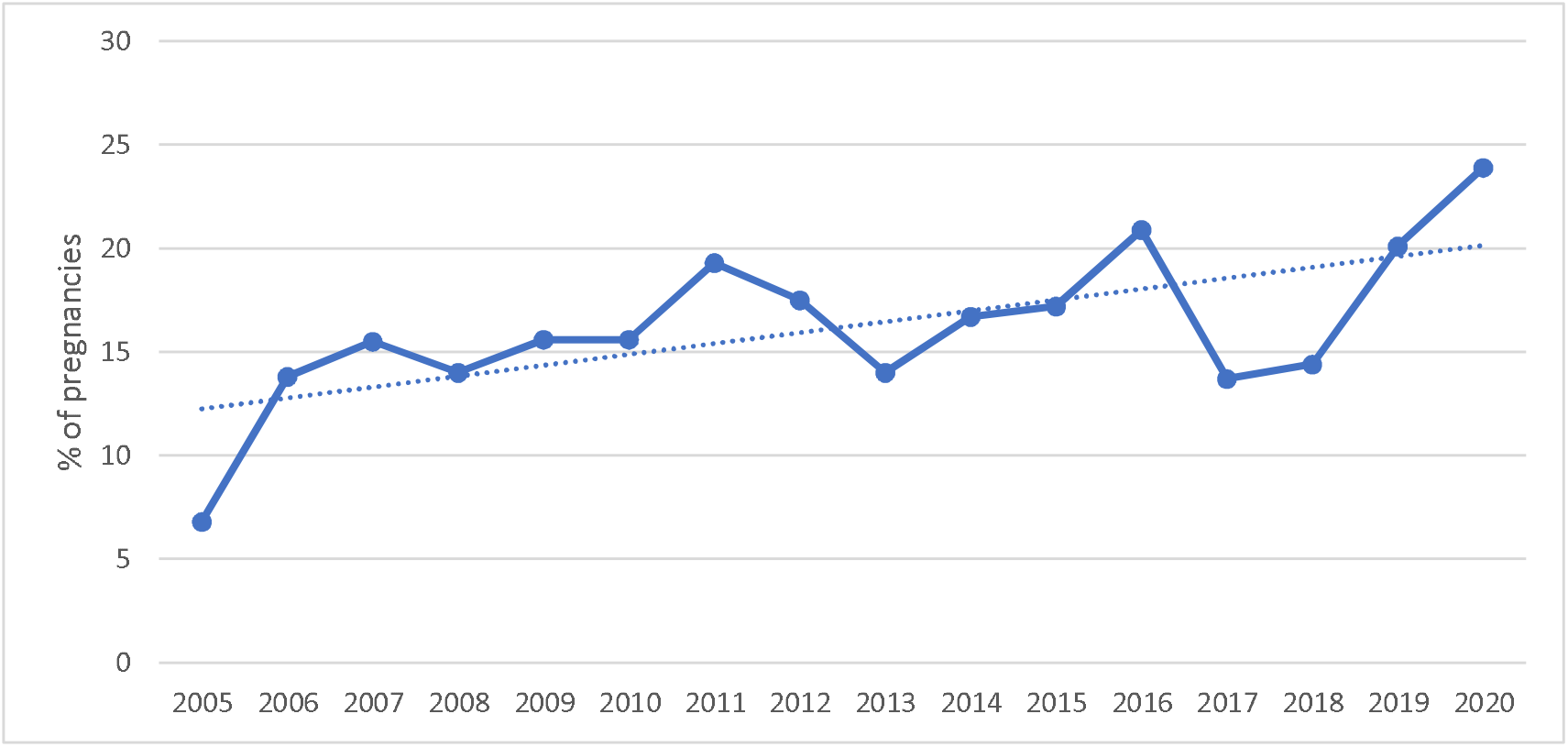
Exposure group “CMZ/MMI at pregnancy onset”: Proportion of pregnancies with a switch to PTU.

#### PTU at pregnancy onset

In 54.9% of the 379 pregnancies exposed to PTU at pregnancy onset, the women had switched before conception from CMZ/MMI to PTU. The proportion of switchers increased from 43.5% in 2005-2008 to 75.0% in 2020. The switch occurred in median 5 (2-12) months before pregnancy onset. In 59.2% of the 379 pregnancies, PTU was also dispensed during pregnancy: in 89.7% of these pregnancies, there was a dispensing during the first trimester, in 45.1% during the second trimester, and in 29.5% during the third trimester. A switch from PTU to CMZ/MMI during pregnancy was observed in 9.0% of pregnancies, in median 15 (13- 19) weeks after pregnancy onset.

#### Start of ATD therapy during pregnancy

In the group of 2,243 pregnancies starting ATD therapy during pregnancy, 1,114 (49.7%) started an ATD therapy with CMZ/MMI during pregnancy and 1,129 (50.3%) started with PTU. The proportion starting CMZ/MMI therapy during pregnancy among all pregnancies in this exposure group decreased from 70.3% in 2005 to 30.8% in 2020 (Fig 6).

**Fig 6.**
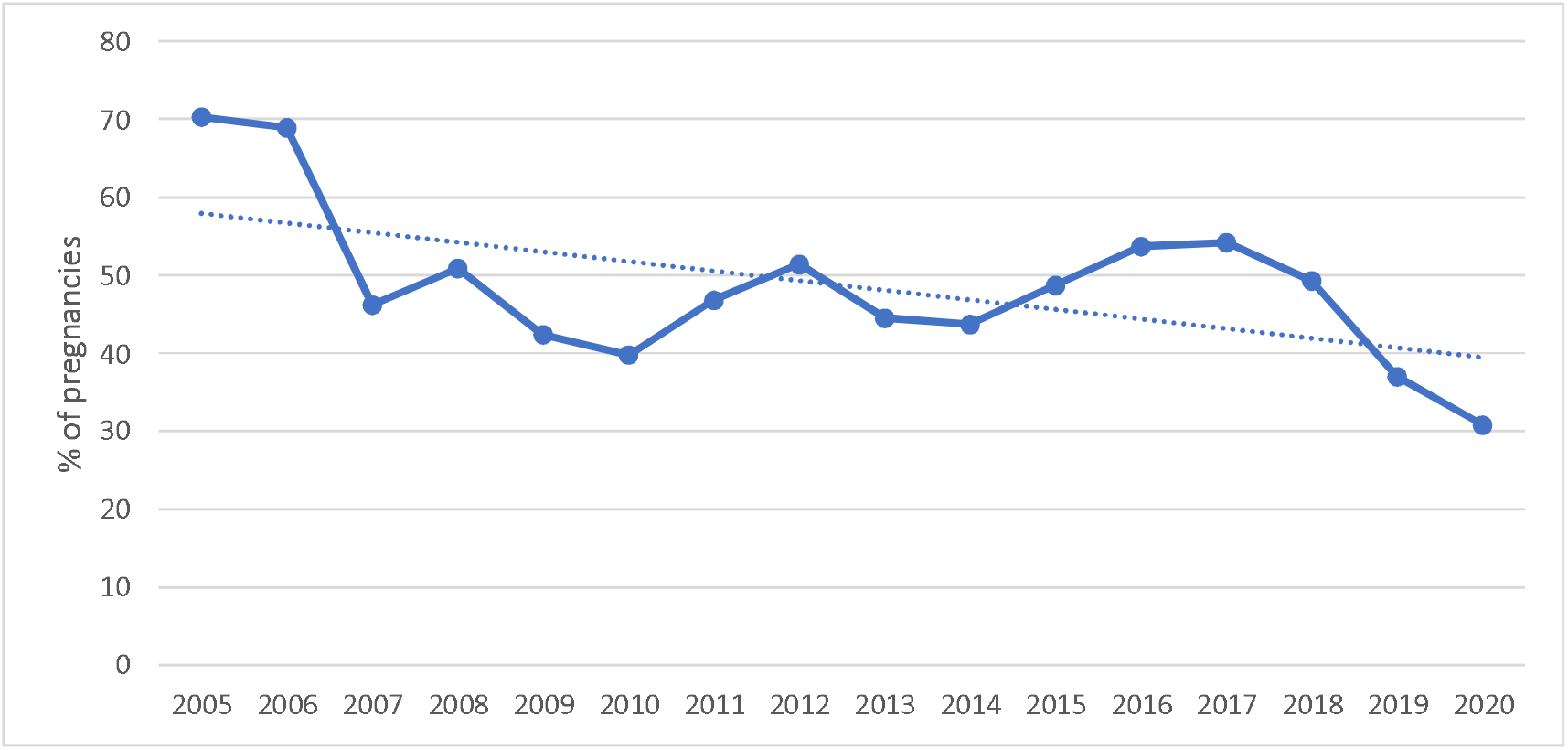
Exposure group “Start of ATD therapy during pregnancy”: Proportion of pregnancies starting with CMZ/TMZ.

In pregnancies starting with CMZ/MMI during pregnancy (n=1,114), the first CMZ/MMI dispensing occurred in median 11 (5-16) weeks after pregnancy onset. In 13.7% of the pregnancies, a switch to PTU was observed, which occurred in median 9 (7-15) weeks after pregnancy onset.

In pregnancies starting with PTU during pregnancy (n=1,129), the first PTU dispensing occurred in median 11 (9-15) weeks after pregnancy onset. In 12.8% of the pregnancies, a switch to CMZ/MMI was observed, which occurred in median 16 (14-20) weeks after pregnancy onset.

## Discussion

In this population-based study covering 20% of the German population, we found that the prescription prevalence of CMZ/MMI in girls and women of childbearing age as well as exposure to CMZ/MMI during the 1^st^ trimester decreased between 2005 and 2020.

The observed decrease in the prescription prevalence of ATDs between 2005 and 2020 among girls and women of childbearing age is consistent with trends in other age groups and males in Germany (9). It indicates a decrease in the prevalence of hyperthyroidism in Germany, supporting the effectiveness of the nationwide iodine fortification program for table salt and cattle food implemented in 1993 in Germany. Introduction of an iodine fortification program may initially increase the prevalence of hyperthyroidism (“iodine-induced hyperthyroidism”), but over time the prevalence of hyperthyroidism usually decreases.(10), i.e. this long-term decreasing effect could be observed during our study period. Similar trends were also observed for the prevalence of relevant iodine-deficient disorders such as goiter and thyroid nodules (11-14). Interestingly, on a substance level, prescription prevalences only decreased for CMZ/MMI, while for PTU they increased until 2019 and remained stable in 2020. This suggests that physicians tend to treat a higher proportion of the (decreasing number of) girls and women of childbearing age with hyperthyroidism with PTU. Given the age pattern observed for the prescription prevalence of PTU (highest prescription prevalence in age group 30-36 years) this trend might be related to an increased awareness of the risks associated with CMZ/MMI in case women become pregnant.

Regarding pregnancies, it is reassuring that the majority of women in our cohort completed ATD therapy before pregnancy. This is in line with the guideline of the European Thyroid Association on the management of Graves’ hyperthyroidism recommending that women with Graves’ hyperthyroidism of reproductive age should be stably euthyroid before attempting pregnancy.(5) Also the decreasing proportion of pregnancies exposed to CMZ/MMI at pregnancy onset, the decreasing proportion of pregnancies with an additional CMZ/MMI dispensing during the first trimester and the increasing proportion of switches from CMZ/MMI to PTU are reassuring. These findings further support that during the study period, prescribers became more and more aware of the potential risks to the unborn child associated with CMZ/MMI. A “Direct Healthcare Professional Communication (DHPC)” emphasizing the risks of CMZ/MMI therapy during pregnancy and reinforcing risk minimization measures in this regard was sent out in February 2019 in Germany (15). The fact that the trends described above already started before 2019 suggests that not only direct healthcare communication but already the increasing evidence and the PRAC referral process increased knowledge and awareness among prescribers regarding the risks of CMZ/MMI during pregnancy.

Nevertheless, among ADT exposed pregnancies, the proportion exposed to CMZ/MMI was still about four times higher than the proportion exposed to PTU, which is the safer alternative during the first trimester. Despite the recommendation to avoid exposure to CMZ/TMZ in the first trimester, there were still 190 pregnancies with estimated exposure to CMZ/TMZ at the beginning of pregnancy and 32 pregnancies in which CMZ/TMZ treatment was started during the first trimester in 2019/2020. Given that GePaRD covers ~20% of the German population, it can be assumed that in the whole of Germany, about 950 pregnancies were still exposed to CMZ/TMZ in 2019/2020. It is not clear whether intolerance to PTU can fully explain this number, i.e. despite the positive developments over time, there might still be room for improvement.

As is the case for all studies based on secondary data, this analysis has limitations inherent to the data source. Information on the prescribed dose and duration is not available in German claims data. We used a pragmatic approach, assuming that all days between two dispensings were exposed if the time between the dispensings was shorter or equal to the time covered by two times the number of DDDs. This may tend to overestimate the length of treatment episodes and thus the number of exposed pregnancies, but from a public health perspective, we prioritized over- rather than underestimating the number of exposed pregnancies. As dosing schemes are not expected to vary across years, we do not think that this uncertainty affects our conclusion regarding the time trends of pregnancies exposed to CMZ/MMI observed in our study. Apart from the uncertainty regarding the prescribed dose, we had no information on actual intake of drugs, as usual in pharmacoepidemiological studies. Women might have stopped ATDs upon learning of their pregnancy, especially in the first weeks. This would also lead to an overestimation of exposed pregnancies.

Strengths of our study are the size and representativeness of the study population, the lack of non-responder or recall bias, and the long study period which allowed us to examine changes over time. We also used validated algorithms for the identification of pregnancies and for estimating pregnancy onset based on the expected delivery date and included pregnancies with no recorded outcome, which are often not considered in database studies, but accounted for 14.2% of the pregnancies.

## Conclusion

The prescription prevalence of ATD, and especially CMZ/MMI, in girls and women of childbearing age decreased between 2005 and 2020. Our study indicates that physicians became increasingly aware of the potential risks of CMZ/MMI as indicated by the decrease in the number of CMZ/MMI dispensings in the 1^st^ trimester, the increase in the number of switches to PTU before and during pregnancy, and the decreasing numbers of starters with CMZ/MMI during pregnancy.

## Data Availability

As we are not the owners of the data we are not legally entitled to grant access to the data of the German Pharmacoepidemiological Research Database. In accordance with German data protection regulations, access to the data is granted only to BIPS employees on the BIPS premises and in the context of approved research projects. Third parties may only access the data in cooperation with BIPS and after signing an agreement for guest researchers at BIPS.

## Acknowledgments

We thank the statutory health insurance providers AOK Bremen/Bremerhaven, DAK- Gesundheit, Techniker Krankenkasse (TK), and hkk Krankenkasse for contributing data for this project and Alina Ludewig and Inga Schaffer for programming the analysis datasets.

## Author Contribution

TS: Conceptualization, Methodology, Validation, Writing – Original Draft, Visualization. BK: Methodology, Software, Validation, Formal Analysis, Writing – Review & Editing. MF: Writing – Review & Editing. UH: Conceptualization, Methodology, Validation, Writing – Original Draft.

## References

1. Nguyen CT, Sasso EB, Barton L, Mestman JH. Graves’ hyperthyroidism in pregnancy: A clinical review. Clin Diabetes Endocrinol. 2018;4:4.

2. Agrawal M, Lewis S, Premawardhana L, Dayan CM, Taylor PN, Okosieme OE. Antithyroid drug therapy in pregnancy and risk of congenital anomalies: Systematic review and meta-analysis. Clin Endocrinol (Oxf). 2022;96(6):857–68.

3. Morales DR, Fonkwen L, Nordeng HME. Antithyroid drug use during pregnancy and the risk of birth defects in offspring: Systematic review and meta-analysis of observational studies with methodological considerations. Br J Clin Pharmacol. 2021;87(10):3890–900.

4. Pharmacovigilance Risk Assessment Committee (PRAC) of the Eurpoean Medicines Agancy. Carbimazole; thiamazole – New information on the known risk of birth defects and neonatal disorders in case of exposure during pregnancy 2019 [Available from: https://www.ema.europa.eu/en/documents/prac-recommendation/prac-recommendations-signals-adopted-26-29-november-2018-prac-meeting_en.pdf.

5. Kahaly GJ, Bartalena L, Hegedüs L, Leenhardt L, Poppe K, Pearce SH. 2018 European Thyroid Association Guideline for the management of Graves’ hyperthyroidism. Eur Thyroid J. 2018;7(4):167–86.

6. Haug U, Schink T. German Pharmacoepidemiological Research Database (GePaRD). In: Sturkenboom M, Schink T, editors. Databases for Pharmacoepidemiological Research. Springer Series on Epidemiology and Public Health: Springer; 2020. p. 119–24.

7. Wentzell N, Schink T, Haug U, Ulrich S, Niemeyer M, Mikolajczyk R. Optimizing an algorithm for the identification and classification of pregnancy outcomes in German claims data. Pharmacoepidemiol Drug Saf. 2018;27(9):1005–10.

8. Schink T, Wentzell N, Dathe K, Onken M, Haug U. Estimating the beginning of pregnancy in German claims data: Development of an algorithm with a focus on the expected delivery date. Front Public Health. 2020;8:350.

9. Thiyagarajan A, Koenen N, Ittermann T, Völzke H, Haug U. Trends in the use of thyroid diagnostics and treatments between 2008 and 2019 in Germany. Sci Rep. 2024;14(1):26710.

10. Zimmermann MB, Boelaert K. Iodine deficiency and thyroid disorders. Lancet Diabetes Endocrinol. 2015;3(4):286–95.

11. Völzke H, Ittermann T, Albers M, Friedrich N, Nauck M, Below H, et al. Five-year change in morphological and functional alterations of the thyroid gland: The Study of Health in Pomerania. Thyroid®. 2012;22(7):737–46.

12. Thamm M, Ellert U, Thierfelder W, Liesenkötter KP, Völzke H. Iodine intake in Germany. Results of iodine monitoring in the German Health Interview and Examination Survey for Children and Adolescents (KiGGS). Bundesgesundheitsblatt Gesundheitsforschung Gesundheitsschutz. 2007;50(5-6):744–9.

13. Khattak RM, Ittermann T, Nauck M, Below H, Völzke H. Monitoring the prevalence of thyroid disorders in the adult population of Northeast Germany. Population Health Metrics. 2016;14(1):39.

14. Manz F, Böhmer T, Gärtner R, Grossklaus R, Klett M, Schneider R. Quantification of iodine supply: Representative data on intake and urinary excretion of iodine from the German population in 1996. Ann Nutr Metab. 2002;46(3-4):128–38.

15. Bundesinstitut für Arzneimittel und Medizinprodukte. Rote Hand Brief (Februar 2019): Carbimazoloder Thiamazol (Synonym: Methimazol)-haltige Arzneimittel - (1) Risiko einer akuten Pankreatitis und (2) Verstärkung der Empfehlung zur Kontrazeption 2019 [Available from: https://www.bfarm.de/SharedDocs/Risikoinformationen/Pharmakovigilanz/DE/RHB/2019/rhb-carbimazol_thiamazol.pdf;jsessionid=F2AE95AF01EEE108994A71F53D7E4323.internet271?blob=publicationFile.

